# Longitudinal changes in home confinement and mental health implications: A 17-month follow-up study in England during the COVID-19 pandemic

**DOI:** 10.1101/2021.10.08.21264749

**Authors:** Feifei Bu, Andrew Steptoe, Daisy Fancourt

## Abstract

**Background:** The COVID-19 pandemic has brought about significant behavioural changes, one of which is increased time spent at home. Although official lockdowns were typically short-term and allowed people to leave their homes for exercise and essential activities, some individuals did not leave their home for prolonged periods due to a range of factors including clinical vulnerability. This study aimed to explore longitudinal patterns of such ‘home confinement’ across different stages of the COVID-19 pandemic in the UK, and its associated predictors and mental health outcomes.

**Methods:** Data were from the UCL COVID -19 Social Study. The analytical sample consisted of 25,390 adults in England who were followed up for 17 months from March 2020 to July 2021. Data were analysed using growth mixture models.

**Results:** Our analyses identified three classes of growth trajectories, including one class showing a high level of persistent home confinement (24.8%), one changing class with clear alignment with national containment policy/advice (32.0%), and one class with a persistently low level of confinement (43.1%). A range of factors were found to be associated the class membership of home confinement trajectories, such as age, gender, income, employment status, social relationships and health. The class with a high level of confinement had the highest number of depressive and anxiety symptoms at the end of the follow-up independent of potential confounders.

**Conclusions:** There was substantial heterogeneity in longitudinal patterns of home confinement during the COVID-19 pandemic. However, a striking proportion of our sample maintained a high level of home confinement over the course of 17 months, even during periods when containment measures were eased or removed and when infection rates were low. They also had the worst mental health outcomes. This group warrants special attention in addressing the mental health impact of the COVID-19 pandemic.

## Introduction

Since December 2019, the world has been devastated by the Coronavirus disease (COVID-19). To control the spread of the disease, in addition to actions such as hand hygiene, face covering and social distancing, lockdowns and ‘stay-at-home’ orders have been carried out in many countries [1]. During lockdowns, people were typically asked to shelter at home, except in circumstances of necessity, with most workplaces, schools and non-essential businesses being closed for extended periods (1 to 3 months). These specific lockdowns suggest on the surface that individuals followed similar patterns of how much time their spent in their homes nationally during the pandemic. However, reports have suggested that in fact individual behaviours varied substantially [2, 3]. First, people at a high risk from COVID-19, such as older adults or those with certain health conditions like lung or heart disease, were advised to remain home even when lockdown measures were eased, and some others followed such heightened precautions themselves voluntarily even in the absence of a specific clinical risk. Second, other individuals did not stay at home even during lockdowns, either due to keyworker roles requiring them to go to work in person, or due to non-compliance. Third, although during lockdowns people were typically allowed to leave their homes for essential reasons such as exercise and shopping for food or medications, reports have suggested that in fact many people did not leave their homes at all each day, thereby engaging in ‘home confinement’. Finally, wide individual variations are expected after lockdowns, with restrictions being eased or as the number of COVID-19 cases changed. However, to date, there has been no research exploring what proportion of people engaged in home confinement and how their patterns of confinement changed across the pandemic, during and after national lockdowns, taking into account potentially heterogeneity in longitudinal changes in home confinement.

‘Home confinement’ is an important behaviour to understand as it could have substantial implications for mental and physical health. Research to date has generally suggested negative health impacts of home confinement. For instance, higher levels of psychological distress [4, 5] and loneliness [6] were found during the COVID-19 lockdown compared to before. It was also found that depressive and anxiety symptoms peaked during lockdown which decreased following the easing of restrictions [7]. Negative changes in eating, drinking and physical activity were also reported, compared to before lockdowns [4, 8, 9]. Potential mechanisms include but not limited to the feeling of infringement upon personal freedom, limited access to health care, reduced social participation and support, related financial loss or other adversities [10, 11]. It is important to note that most of existing evidence come from studies comparing lockdowns to non-lockdown periods. However, as discussed above, although lockdown limits how much people could leave their homes but does allow people to go outside each day, it is not the same as home confinement. However, there has been little research on the health implications of stricter behaviour of home confinement more specifically.

Further, there has been limited research so far into who demographically were more likely to engage in home confinement during the COVID-19 pandemic. Recent evidence on ‘compliance’ showed that younger age, male gender, higher educational level and living alone were associated with lower levels of compliance [12]. Another study found that age, gender, education, employment status, living alone, physical and mental health, personality were related to the growth trajectory of compliance [13]. However, there is a substantial distinction between ‘compliance’ and ‘home confinement’. First, the COVID-19 rules and guidelines were broad and included hand hygiene, face covering, social distancing and so forth, so using ‘compliance’ as a proxy for understanding home confinement is flawed. Moreover, as going outdoors was not prohibited even during strict lockdown periods in England as long as it was for essential reasons such as exercise, home confinement may in fact be only weakly related to compliance. Indeed, leaving one’s home for exercise was still recommended for maintaining health, so one could still fully comply with the lockdown ‘stay-at-home’ orders but nonetheless leave the home every day. Given the possible negative health effects of home confinement, it is important to identify who engaged in this behaviour so that resources can be targeted to support these individuals if needed.

Therefore, this study examined home confinement defined as staying at home without leaving the property (except for onto a balcony or into a garden). We used data from 25,390 adults living in England who were followed for 17 months between March 2020 and July 2021 to model longitudinal changes in home confinement. We used growth mixture modelling approach allowing for different patterns of growth trajectory. Further, this study sought to explore factors that might be associated with different patterns of longitudinal changes in home confinement and their mental health implications. This will facilitate a better understanding of behavioural response in longer term and its predictors in the face of public health crisis.

## Methods

### Study design and participants

This study analysed data from the University College London (UCL) COVID-19 Social Study, a large panel study of the psychological and social experiences of over 75,000 adults (aged 18+) in the UK during the COVID-19 pandemic. The study commenced on 21 March 2020 and involves weekly and then monthly (four-weekly) online data collection from participants for the duration of the pandemic. The study did not use a random sample design and therefore the original sample is not representative of the UK population. However, it does contain a heterogeneous sample that was recruited using three primary approaches. First, convenience sampling was used, including promoting the study through existing networks and mailing lists (including large databases of adults who had previously consented to be involved in health research across the UK), print and digital media coverage, and social media. Second, more targeted recruitment was undertaken focusing on (i) individuals from a low-income background, (ii) individuals with no or few educational qualifications, and (iii) individuals who were unemployed. Third, the study was promoted via partnerships with third sector organisations to vulnerable groups, including adults with pre-existing mental health conditions, older adults, carers, and people experiencing domestic violence or abuse. The study was approved by the UCL Research Ethics Committee [12467/005] and all participants gave informed consent. A full protocol for the study is available online at https://github.com/UCL-BSH/CSSUserGuide.

In the present study, we restricted the sample to participants living in England (N= 58,486). Further, we included only participants with at least three repeated measures between 21 March 2020 and 25 July 2021. These criteria provided us with data from 37,800 participants. Around 10% of these participants had missing data for demographic, social and health-related predictors and another 3.1% on mental health measures. After excluded these participants, we had sample size of 32,856 participants. Finally, we excluded keyworkers (people working in essential sectors such as health and social care, transport etc.) who were in employment, given they were likely to have an established pattern of home confinement especially during lockdown periods. This provided us with a final analytical sample of 25,390 participants who were followed up for a maximum of 17 months.

### Measures

#### Home confinement

Participants were asked every week: ‘In the past 7 days, how many days have you not left the house or garden?’ The options ranged from 0 to 7 days. This was analysed as a continuous variable.

#### Predictors

We considered a range of socio-demographic, social, health and psychological factors as potential predictors. These included gender (women vs men), ethnicity (white vs ethnic minorities), age groups (age 18-29, 30-45, 46-59, 60+), education (up to GCSE levels, A-levels or equivalent, and university degree or above), income (<£30,000 vs ≥£30,000 per annum), employment status (employed vs other), area of living (rural vs urban) and dog ownership (yes vs no). Social factors included living situation (living alone vs living with others), number of close friends (0 to 10+) and usual social contacts (1 to 5, from less than once a month to every day). In addition, we included two health-related factors: self-reported diagnosis of any long-term physical health condition (e.g., asthma or diabetes) or any disability (yes vs no), and self-reported diagnosis of any long-term mental health condition (e.g., depression, anxiety) (yes vs no). Psychological factors included Big Five personality traits (neuroticism, extraversion, openness, agreeableness and conscientiousness)[14] and COVID-19 stress. The latter was measured by asking participants if they had experienced any minor/major stress about catching COVID-19 or becoming serious ill from it. This was coded as none, minor stress and major stress.

#### Distal outcomes

To explore mental health implications of home confinement, we looked at depressive and anxiety symptoms. Depressive symptoms were measured using the Patient Health Questionnaire (PHQ-9) [15]; a standardised instrument for screening for depression in primary care. Unlike the original PHQ-9, the current study enquired about symptoms ‘over the last week’ instead of ‘over the last two weeks’ as data were initially collected weekly. The questionnaire includes 9 items with 4-point responses ranging from ‘not at all’ to ‘nearly every day’. Higher overall scores indicate more depressive symptoms, ranging from 0 to 27. Anxiety symptoms were measured using the Generalized Anxiety Disorder assessment (GAD-7) [16]; a well-validated tool used to screen for generalized anxiety disorder in clinical practice and research. These questions were also worded as ‘over the last week’ for the same reason as the depression items. The GAD-7 comprises 7 items with 4-point responses ranging from ‘not at all’ to ‘nearly every day’, with higher overall scores indicating more symptoms of anxiety, ranging from 0 to 21. Both depressive and anxiety symptoms were measured longitudinally. We used measures at the last time point when participants were observed as distal outcomes, while controlling for the relevant mental health measures at the baseline.

### Statistical analysis

Data were analysed using the growth mixture modelling (GMM) approach. The conventional growth modelling approach assumes one homogeneous growth trajectory, allowing individual growth factors to vary randomly around the overall mean. GMM relaxes this assumption and enables researchers to explore different patterns of change (latent trajectory classes) [17]. More specifically, we used GMM with free time scores, in which time scores (months) were estimated as free parameters, except for two fixed at 0 and 1 for the purpose of model identification. In this model, we made no assumption about the shape of growth trajectories which was left to be determined by data.

Starting with the unconditional GMM, we compared models with different number of classes on the basis of the Bayesian information criterion (BIC) and sample-size adjusted Bayesian information criterion (ABIC), along with the Vuong-Lo-Mendell-Rubin likelihood ratio (LMR-LR) test and Adjusted Lo-Mendell-Rubin likelihood ratio (ALMR-LR) test. After identifying the optimal number of classes, we introduced covariates to the model to explore what factors were associated with class membership, which was fitted using logistic regression model. Next, we included mental health measures distal outcomes, with depressive and anxiety symptoms being modelled separately controlling for baseline mental health measures (see Figure S2 in the Supplement for model specification). This allowed us to examine how latent classes were related to mental health at the end of the follow-up. These models were fitted using the new three-step approach, which accounts for measurement errors in class predictions [18, 19]. One-step conditional GMM models were fitted as sensitivity analyses. Weights were applied throughout the analyses. The final analytical sample was weighted to the proportions of gender, age, ethnicity and education in the English population obtained from the Office for National Statistics [20]. The main analyses were implemented in Mplus Version 8.

## Results

### Sample characteristics

Before weighting the 25,390 participants, there was an over-representation of women and people with a degree or above and under-representation of people from ethnic minority backgrounds and adults under 30 (Table 1). After weighting, the sample reflected population proportions, with 50.7% women, 34.6% with a degree or above, 12.3% of ethnic minority and 17.9% under 30.

**Table 1.** Descriptive statistics of the sample before and after weighting (N=25,390)

|  |  | Raw data |  | Weighted data |  |
| --- | --- | --- | --- | --- | --- |
|  |  | %/Mean (SD) | N | %/Mean (SD) | N |
| Gender | Women | 74.1% | 18,807 | 50.7% | 12,714 |
|  | Men | 25.9% | 6,583 | 49.3% | 12,342 |
| Ethnicity | Minority | 4.5% | 1,152 | 12.3% | 3,090 |
|  | White | 95.5% | 24,238 | 87.7% | 21,966 |
| Age | 18-29 | 6.3% | 1,607 | 17.9% | 4,486 |
|  | 30-45 | 24.6% | 6,250 | 24.2% | 6,064 |
|  | 46-59 | 30.3% | 7,699 | 22.0% | 5,516 |
|  | 60+ | 38.7% | 9,834 | 35.9% | 8,990 |
| Education | Low (GCSEs or below) | 13.6% | 3,463 | 33.6% | 8,424 |
|  | Medium (A-levels or equivalent) | 16.8% | 4,275 | 31.8% | 7,966 |
|  | High (Degree or above) | 69.5% | 17,652 | 34.6% | 8,666 |
| Household income (annual) | <30k | 40.4% | 10,269 | 49.3% | 12,342 |
|  | ≥30k | 59.6% | 15,121 | 50.7% | 12,714 |
| Employment status | Employed | 54.5% | 13,833 | 48.5% | 12,148 |
|  | Other | 45.5% | 11,557 | 51.5% | 12,908 |
| Living status | Living alone | 21.7% | 5,514 | 19.8% | 4,961 |
|  | Living with others | 78.3% | 19,876 | 80.2% | 20,095 |
| Area of living | Urban | 78.2% | 19,865 | 79.7% | 19,980 |
|  | Rural | 21.8% | 5,525 | 20.3% | 5,076 |
| Dog ownership | Yes | 22.3% | 5,651 | 23.4% | 5,863 |
|  | No | 77.7% | 19,739 | 76.6% | 19,193 |
| Number of close friends (range: 0-10) |  | 4.9 (3.1) | 25,390 | 4.5 (3.1) | 25,056 |
| Usual social frequency (range: 1-5) |  | 3.1 (1.1) | 25,390 | 3.0 (1.2) | 25,056 |
| Physical health condition | Yes | 39.4% | 9,996 | 39.9% | 9,995 |
|  | No | 60.6% | 15,394 | 60.1% | 15,061 |
| Mental health condition | Yes | 17.8% | 4,518 | 20.6% | 5,150 |
|  | No | 82.2% | 20,872 | 79.4% | 19,906 |
| Big Five personality (range: 3-21) | Neuroticism | 11.3 (4.3) | 25,390 | 11.5 (4.5) | 25,056 |
|  | Extraversion | 12.9 (4.3) | 25,390 | 12.6 (4.3) | 25,056 |
|  | Openness | 15.5 (3.3) | 25,390 | 15.0 (3.3) | 25,056 |
|  | Agreeableness | 15.5 (3.1) | 25,390 | 15.3 (3.2) | 25,056 |
|  | Conscientiousness | 15.8 (3.0) | 25,390 | 15.5 (3.1) | 25,056 |
| COVID-19 stress | None | 40.0% | 10,155 | 42.5% | 10,648 |
|  | Minor stress | 37.2% | 9,452 | 34.0% | 8,528 |
|  | Major stress | 22.8% | 5,783 | 23.5% | 5,880 |
| Depressive symptoms baseline (range: 0-27) |  | 6.4 (5.8) | 25,390 | 7.0 (6.3) | 25,056 |
| Depressive symptoms last wave (range: 0-27) |  | 5.6 (5.8) | 25,390 | 6.1 (6.3) | 25,056 |
| Anxiety symptoms baseline (range: 0-21) |  | 5.3 (5.2) | 25,390 | 5.6 (5.6) | 25,056 |
| Anxiety symptoms last wave (range: 0-21) |  | 4.2 (5.1) | 25,390 | 4.6 (5.5) | 25,056 |

### Latent trajectory classes

To determine the optimal number of latent trajectory classes, we compared across unconditional GMMs with different numbers of classes. Although the BIC and ABIC continued to decrease with each additional class being added to the model, the ALMR-LR test of the four-class GMM did not reject the three-class model (Table S2). Therefore, the three-class GMM model was chosen. It had an adequate quality of class membership classification (entropy=0.66). Figure 1 shows the estimated longitudinal growth trajectory of home confinement for each latent class (LC). The Figure also includes the Stringency Index of the strictness of lockdown policies in England, with higher scores indicating greater strictness [1], and the number of new cases of COVID-19 infection according to the Government of the United Kingdom [21].

**Figure 1.**
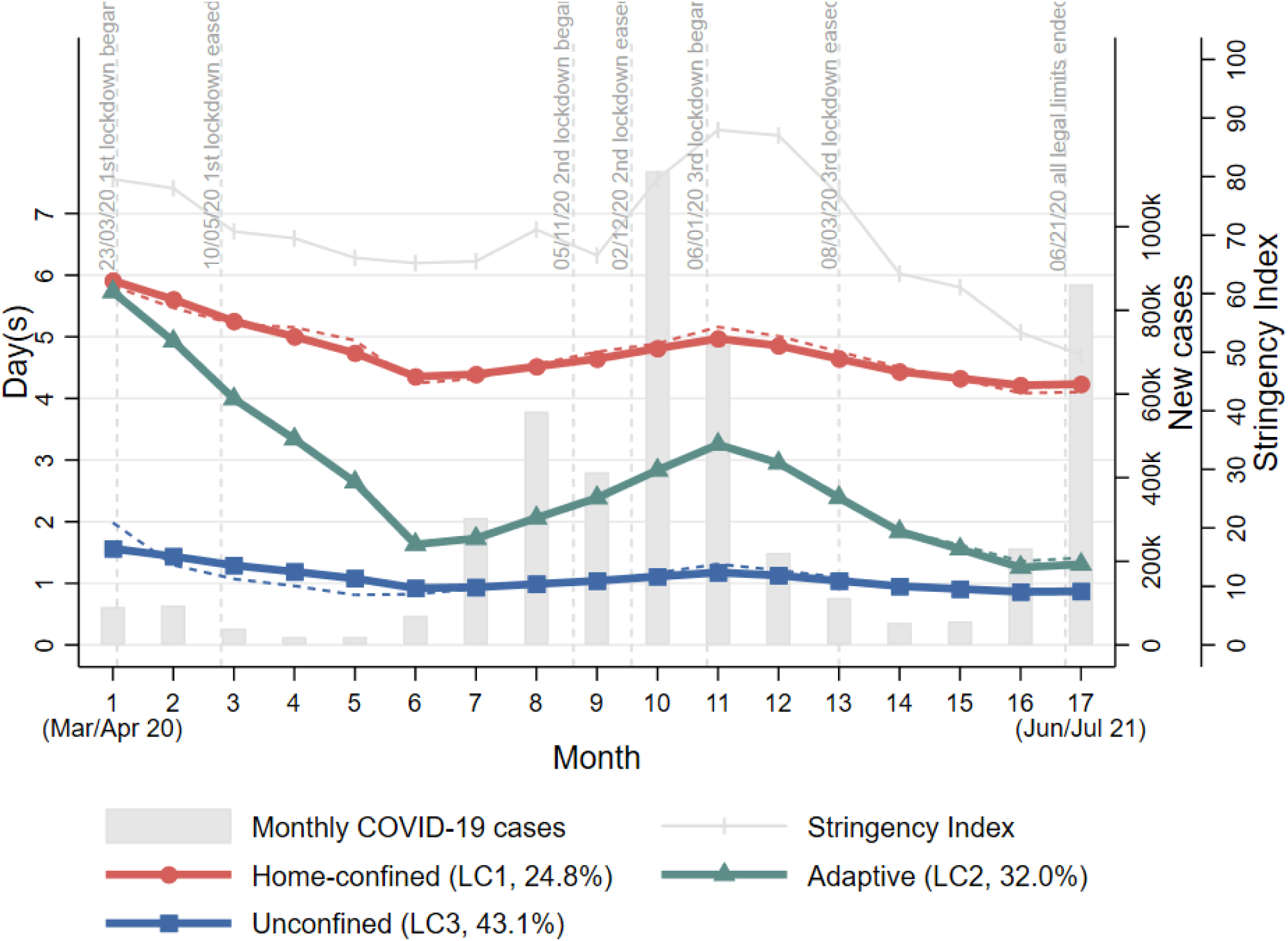
Estimated growth trajectories for different classes of home confinement (March 2020-July 2021)

**Figure 1.**
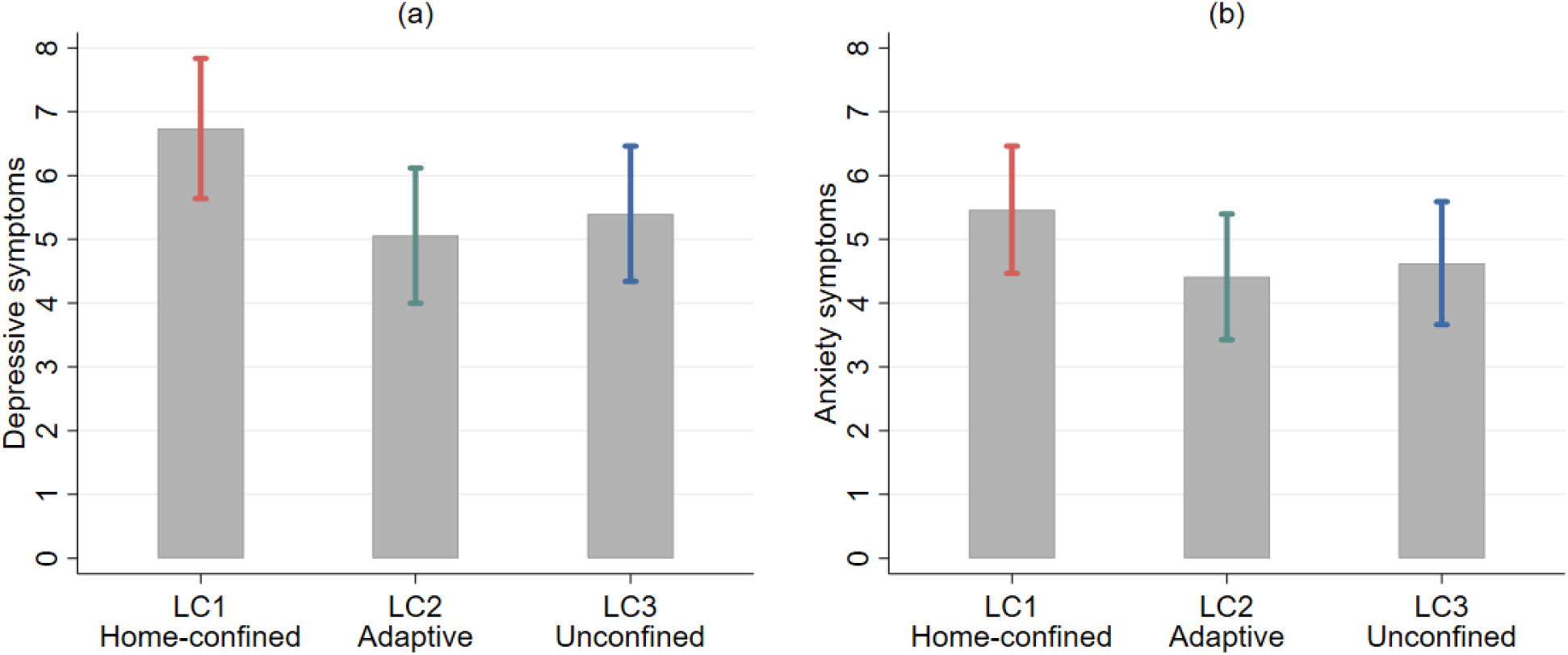
Estimated depressive and anxiety symptoms at the end of the follow-up by class membership (Notes: Categorical predictors were set to use the reference and continuous variables to the mean, additionally controlling for depressive/anxiety measure at baseline respectively.)

The first class (24.8%) was marked by a high level of home confinement (4 to 6 days a week) persisting throughout the 17-month follow-up period, even when lockdown measures were eased with stringency index being relatively low. Therefore, it was labelled as the ‘home-confined’. However, this class did show a decline in days in confinement between March and August 2020 before it started to increase in September 2020 and then decrease again around February 2021. This trend was in line with the changes in national policies and to a lesser extent with the number of COVID-19 cases with numbers being relatively low at the start (Figure 1). The second class (32.0%) started off with a high level of confinement at the beginning of the first national lockdown, which declined sharply (from 6 to 1.7 days a week) into summer months. This was followed by an increase from September 2020 and peaked in January 2021 when the third national lockdown started, before it started to decrease again. This class was labelled as the ‘adaptive’ who seemed to adapt their behaviours to circumstance changes to a much greater extent compared to the ‘home-confined’. The third class was the largest (43.1%), which consisted of people showing a very low level of home confinement consistently (about 1 day a week) regardless of policy changes and COVID-19 cases. They were regarded as the ‘unconfined’.

### Factors associated with latent trajectory classes

We fitted a logistic regression model to examine how individual characteristics were related to class membership of home confinement trajectories, using LC1 (‘home-confined’) as the reference (Table 2). Women had 24% lower odds of being adaptive (LC2) and 42% lower odds of being unconfined (LC3) than men. There was no ethnic difference comparing LC2 to LC1, but people from ethnic minority backgrounds had 35% lower odds of being unconfined (LC3) relative to being home-confined. People aged 46 or above had lower odds of being adaptive (OR=0.55-0.56), but older adults aged 60 or above had 54% high odds of being in the unconfined class than young adults aged 18 to 29. There was no educational difference comparing LC2 to LC1, but those with a higher education qualification had 57% higher odds of being unconfined. People from low-income households had 24% lower odds of being adaptive and 36% lower odds of being unconfined. People who were employed at the baseline had 2.4 times odds of being adaptive and 2.42 times odds of being unconfined than those who were not in employment. There was no evidence that rurality was related to latent trajectory classes. There was no evidence that dog ownership was related to the odds of being adaptive, but people who owned a dog had 2.64 times odds of being unconfined.

**Table 2.**
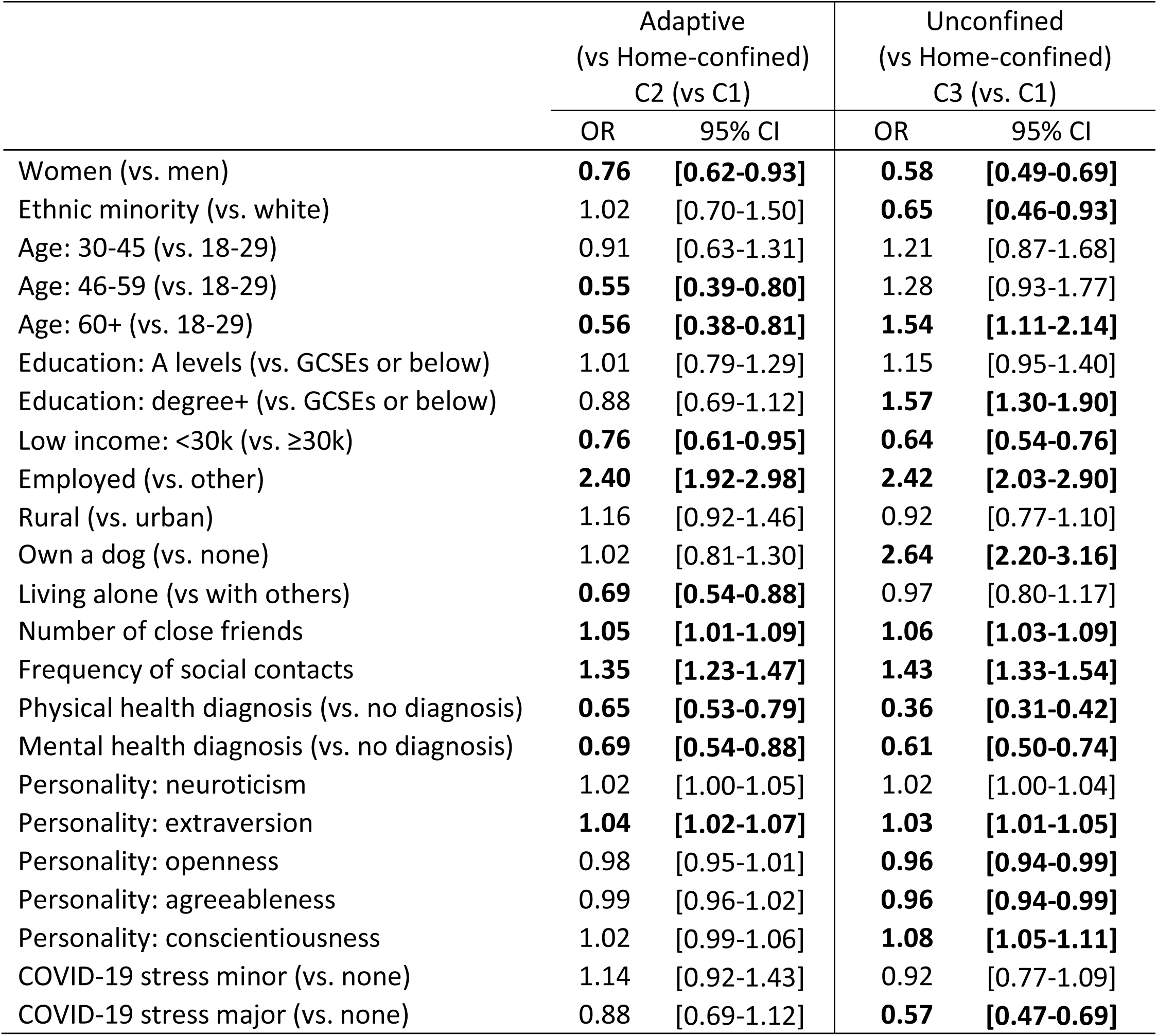
Results from the 3^rd^ step logistic regression model (N=25,390)

|  | Adaptive<br>(vs Home-confined)<br>C2 (vs C1) |  | Unconfined<br>(vs Home-confined)<br>C3 (vs. C1) |  |
| --- | --- | --- | --- | --- |
|  | OR | 95% CI | OR | 95% CI |
| Women (vs. men) | **0.76** | **[0.62-0.93]** | **0.58** | **[0.49-0.69]** |
| Ethnic minority (vs. white) | 1.02 | [0.70-1.50] | **0.65** | **[0.46-0.93]** |
| Age: 30-45 (vs. 18-29) | 0.91 | [0.63-1.31] | 1.21 | [0.87-1.68] |
| Age: 46-59 (vs. 18-29) | **0.55** | **[0.39-0.80]** | 1.28 | [0.93-1.77] |
| Age: 60+ (vs. 18-29) | **0.56** | **[0.38-0.81]** | **1.54** | **[1.11-2.14]** |
| Education: A levels (vs. GCSEs or below) | 1.01 | [0.79-1.29] | 1.15 | [0.95-1.40] |
| Education: degree+ (vs. GCSEs or below) | 0.88 | [0.69-1.12] | **1.57** | **[1.30-1.90]** |
| Low income: <30k (vs. ≥30k) | **0.76** | **[0.61-0.95]** | **0.64** | **[0.54-0.76]** |
| Employed (vs. other) | **2.40** | **[1.92-2.98]** | **2.42** | **[2.03-2.90]** |
| Rural (vs. urban) | 1.16 | [0.92-1.46] | 0.92 | [0.77-1.10] |
| Own a dog (vs. none) | 1.02 | [0.81-1.30] | **2.64** | **[2.20-3.16]** |
| Living alone (vs with others) | **0.69** | **[0.54-0.88]** | 0.97 | [0.80-1.17] |
| Number of close friends | **1.05** | **[1.01-1.09]** | **1.06** | **[1.03-1.09]** |
| Frequency of social contacts | **1.35** | **[1.23-1.47]** | **1.43** | **[1.33-1.54]** |
| Physical health diagnosis (vs. no diagnosis) | **0.65** | **[0.53-0.79]** | **0.36** | **[0.31-0.42]** |
| Mental health diagnosis (vs. no diagnosis) | **0.69** | **[0.54-0.88]** | **0.61** | **[0.50-0.74]** |
| Personality: neuroticism | 1.02 | [1.00-1.05] | 1.02 | [1.00-1.04] |
| Personality: extraversion | **1.04** | **[1.02-1.07]** | **1.03** | **[1.01-1.05]** |
| Personality: openness | 0.98 | [0.95-1.01] | **0.96** | **[0.94-0.99]** |
| Personality: agreeableness | 0.99 | [0.96-1.02] | **0.96** | **[0.94-0.99]** |
| Personality: conscientiousness | 1.02 | [0.99-1.06] | **1.08** | **[1.05-1.11]** |
| COVID-19 stress minor (vs. none) | 1.14 | [0.92-1.43] | 0.92 | [0.77-1.09] |
| COVID-19 stress major (vs. none) | 0.88 | [0.69-1.12] | **0.57** | **[0.47-0.69]** |

All social factors were found to be associated with latent trajectory classes. People living alone had 31% lower odds of being adaptive, but no difference was found comparing LC3 to LC1. People with more close friends had higher odds of being both adaptive (OR=1.05, 95% CI=1.01-1.09) and unconfined (OR=1.06, 95% CI=1.03-1.09). Similarly, frequent social contacts were also associated with higher odds of being adaptive (OR=1.35, 95% CI=1.23-1.47) and unconfined (OR=1.43, 95% CI=1.33-1.54).

People with pre-existing physical health conditions had 35% lower odds of being in the adaptive class and 64% lower odds in the unconfined class. Similarly, people with mental health conditions had 31% lower odds of being adaptive and 39% lower odds of being unconfined. Personality trait, extraversion was associated with higher odds of being adaptive (OR=1.04, 95% CI=1.02-1.07). Extraversion and conscientiousness were also associated with higher odds of being unconfined (OR=1.03-1.08); whereas openness and agreeableness were associated with lower odds of being unconfined (OR=0.96). Finally, there was little evidence that stress was related to latent trajectory classes, except that having major stress related to COVID-19 at the baseline were associated with lower odds of being unconfined (OR=0.57, 95% CI=0.47-0.69).

### Mental health by latent trajectory classes

Models including mental health measures as distal outcomes addressed the question whether the patterns of home confinement trajectories were related to depressive and anxiety symptoms. The estimated depressive and anxiety symptoms and their 95% confidence intervals by latent classes were presented in Figure 2. People in the home-confined classed had the highest number of depressive symptoms independent of all covariates and depressive symptoms at baseline, followed by the unconfined. People from the adaptive class had the lowest number of depressive symptoms. All class differences were statistically significant (LC1-LC2: diff=1.68, p<0.001; LC1-LC3: diff=1.34, p<0.001; LC3-LC2: diff=0.34, p=0.025). Similar results were also found for anxiety symptoms. The home-confined had higher levels of anxiety than the adaptive (diff=1.05, p<0.001) and unconfined (diff=0.84, p<0.001). However, the difference between the adaptive and unconfined was not statistically significant (diff=0.21, p=0.150).

## Discussion

This study is the first to examine the longitudinal changes in home confinement during the COVID-19 pandemic. Using data from England, our analyses identified three unique classes of growth trajectories of home confinement. About one in four participants had a high level of confinement persistently across the 17-month follow-up between March 2020 and July 2021. Nearly a third participants started with a high level of confinement which decreased sharply into summer months, then rose and fell in accordance to changes in containment measures. The largest proportion of participants (43%) had little change over time, showing a very low level of home confinement.

The class membership was related to a range of socio-demographic, social, health and psychological factors. Most likely to be in the class of high-levels of home confinement were those with pre-existing physical health conditions. This was as expected given guidance from the World Health Organization (WHO) and National Health Service (NHS) advising people with specific health conditions to shield. Notably these people were reported having poorer psychosocial experience during the pandemic [22, 23], which might be partially explained by their high level of home confinement. People with pre-existing mental health conditions were also more likely to engage in persistent home confinement, but interestingly neither neuroticism nor being stressed about catching or becoming ill from COVID-19 was not related to engaging in home confinement rather than simply following the guidelines (‘adaptive’ class). This suggests two things. First, concern about the virus itself did not lead people to impose stricter rules on their own behaviours than the national guidance (although people who were majorly stressed about COVID-19 were less likely to be in the ‘unconfined’ class). Second, pandemic-related stress itself or general neurotic traits did not lead people to confine themselves to their homes, but rather pre-existing mental illness was the specific predictor. This is consistent with findings from a qualitative study showing that leaving the house was a cause of anxiety which led to prolonged home confinement among adults with mental health conditions [24].

Amongst other predictors, women were more likely to engage in home-confinement than men. This is consistent with findings that women reported higher level of compliance to containment rules and guidelines [12, 13], and were more likely to self-isolate after developing symptoms of COVID-19 [25]. Recent studies generally reported a higher level of psychological distress among women during the COVID-19 pandemic [23, 26], so it is possible that the higher level of home confinement amongst women as found in our study partly explain these prior findings. In considering why women more than men engaged in home confinement, our findings were independent of other factors that could plausibly explain this relationship such as stress and mental health (both higher in women during the pandemic). But one explanation could lie in the division of labour: women were found to spend much more time on unpaid care and more likely to reduce working hours to accommodate increasing childcare demand during lockdowns [26]. This may have reduced available time to go outdoors for fresh air or exercise. Our findings for age were complex. Adults aged 60+ were more likely to be the classes of home-confined and unconfined. This suggests a heterogeneity in older people’s responses to COVID-19, and highlights that there was more nuance in older adults’ behaviours than merely the higher compliance among older people often reported [12, 13]. Notably the two classes that were associated with older age showed relatively little change over time. With increasing awareness of the importance of maintaining mental wellbeing during COVID-19 [27, 28], it is possible that older adults were keen to stick to their existing routines, using going out as a means to be physically active and to cope with mental health challenges. A qualitative study reported that some older adults focused more on their health during the COVID-19 pandemic and became physically more active by going for regular walks and taking up new forms of physical activity [29].

Another factor that was associated with higher odds of being home-confined persistently is low household income. As we only controlled for employment status at the baseline, it is probable that people from low-income households were likely to be in jobs that were more subject to unemployment and furlough schemes during the follow-up [30, 31], which could contribute to why they were less likely to have daily reasons to leave the house to go to work. But it does not fully explain why their home confinement would be persistently higher.

Social factors were also found to predict class membership. Adults who lived alone were more likely to engage in home-confinement (relative to being adaptive) than those living with others, although there was no difference when comparing the home-confined to unconfined. Small size of friend network and less frequent social contacts were also associated with higher odds of persistently engaging in home confinement comparing to either the adaptive or unconfined class. These findings suggest that people who were socially more isolated at the start of the pandemic were at risk of a high level of home-confinement throughout the pandemic, which in turn might have exacerbated their sense of social isolation. These individuals were not merely less extrovert, as although adults who scored lower on extraversion were consistently more likely to be home-confined, our analyses simultaneously controlled for extraversion, so existing social isolation remained an independent risk factor. Instead, the mechanism may be related to a lack of social support and motivation, and unhealthy lifestyles, such as lower physical activity and more sedentary behaviours as suggested in previous literature [32, 33]. This might be exacerbated during the COVID-19 pandemic when usual social and daily routines were descripted.

Further to predictors of latent trajectory classes, the study addresses another important question about whether patterns of home-confinement trajectories are related to mental health outcomes. Our analyses revealed a much higher number of depressive and anxiety symptoms among people who engaged in home confinement persistently. This is in line with well-established evidence of the negative mental health impact of social isolation in general prior to the COVID-19 pandemic [34–36], and recent evidence linking lockdowns with a range of negative outcomes such as domestic violence [37, 38], physical inactivity [8], weight gain [39], alcohol consumption [9] and psychological strains, such as loneliness, life satisfaction, depressive symptoms [5, 11, 22]. However, it is worthy of note that people in the unconfined class had more depressive symptoms than those who were adaptive. Similar finding was also found for anxiety symptoms albeit statistically insignificant. One possible explanation is that for some going out of the house was not necessarily by choice but for practical reasons, for example work or other obligations such as caring (even though we excluded keyworkers from the analysis). In other words, it is possible that some people in the unconfined class were unable rather than unwilling to reduce their days of going out, especially during national lockdowns. Although going outdoors comes with mental health benefits in general, this aspect of external obligations and the associated health risks may have had a negative impact on people’s mental health, which may explain the difference in depressive symptoms between the unconfined and adaptive.

This study has a number of strengths including its large sample size, repeated weekly follow-up of the same participants over 17 months since the first UK lockdown across three national lockdown periods, and robust statistical approaches. Although the UCL COVID-19 Social Study did not use a random sample, it does have a large sample size with wide heterogeneity, including good stratification across all major socio-demographic groups. In addition, analyses were weighted on the basis of population estimates of core demographics, with the weighted data showing good alignment with national population statistics and another large scale nationally representative social survey [40]. Despite all efforts to make our sample inclusive and representative of the adult population in England, we cannot rule out the possibility of potential biases due to omitting other demographic factors that could be associated with survey participation in the weighting process. It is also important to acknowledge that our data were collected through online surveys, so people without internet access were excluded. It is possible that these people may be more likely to be home confined, in which case, the level of home confinement could be slightly underestimated. Further, there is a lack of pre-pandemic information. Therefore, it is unclear how the growth trajectories of home confinement are related to patterns before the pandemic, and it is possible that some of the individuals in the home-confined category habitually spend more of their time at home. Future studies could extend our analyses to explore whether, and to what extent, the longitudinal patterns of home confinement persist after the COVID-19 pandemic.

Lockdowns and stay-at-home orders have been shown to be essential and effective in controlling the COVID-19 outbreak, but there have been concerns about their potential negative impacts on the mental health of the public [10, 41]. Our study showed that a large proportion of adults went out almost everyday during and after lockdowns, and about a third adjusted their days in home confinement in according to policy changes. However, one in four adults maintained a high level of home confinement throughout lockdowns and across the seasons, even during periods when containment measures were eased or removed and when infection rates were low. The groups at a higher risk of persistent home-confinement were women and those with low household income, more limited social relationships, and physical or mental health conditions. These findings are concerning as our analyses have shown that persistent home confinement is related to worse mental health outcomes. It is promising that the mental health impact of the COVID-19 pandemic is well acknowledged and the Government of the UK has set out an action plan during 2021 to 2022 to address these issues [42]. Based on findings from this study, we advocate that persistent home confinement should be given special attention in future action plan and interventions. In addition to services for the general public, targeted interventions for at-risk groups (aforementioned) are also needed.

## Data Availability

Anonymous data will be made publicly available following the end of the pandemic.

## Declarations

### Ethics approval and consent to participate

The study was approved by the UCL Research Ethics Committee [12467/005] and all participants gave informed consent.

### Availability of data and materials

Anonymous data will be made publicly available following the end of the pandemic.

### Competing interests

All authors declare no conflicts of interest.

### Funding

This Covid-19 Social Study was funded by the Nuffield Foundation [WEL/FR-000022583], but the views expressed are those of the authors and not necessarily the Foundation. The study was also supported by the MARCH Mental Health Network funded by the Cross-Disciplinary Mental Health Network Plus initiative supported by UK Research and Innovation [ES/S002588/1], and by the Wellcome Trust [221400/Z/20/Z]. DF was funded by the Wellcome Trust [205407/Z/16/Z].

### Author contributions

FB, AS and DF developed the study idea and the analysis plan. FB analysed the data and wrote the first draft. All authors had accessed and verified the underlying data. All authors provided critical revisions, read and approved the submitted manuscript.

## Acknowledgements

The researchers are grateful for the support of a number of organisations with their recruitment efforts including: the UKRI Mental Health Networks, Find Out Now, UCL BioResource, SEO Works, FieldworkHub, and Optimal Workshop. The study was also supported by HealthWise Wales, the Health and Care Research Wales initiative, which is led by Cardiff University in collaboration with SAIL, Swansea University.

## Notes

### Competing Interest Statement

The authors have declared no competing interest.

