## Supplement for "Longitudinal changes in home confinement and mental health implications: A 17-month follow-up study in England during the COVID-19 pandemic"

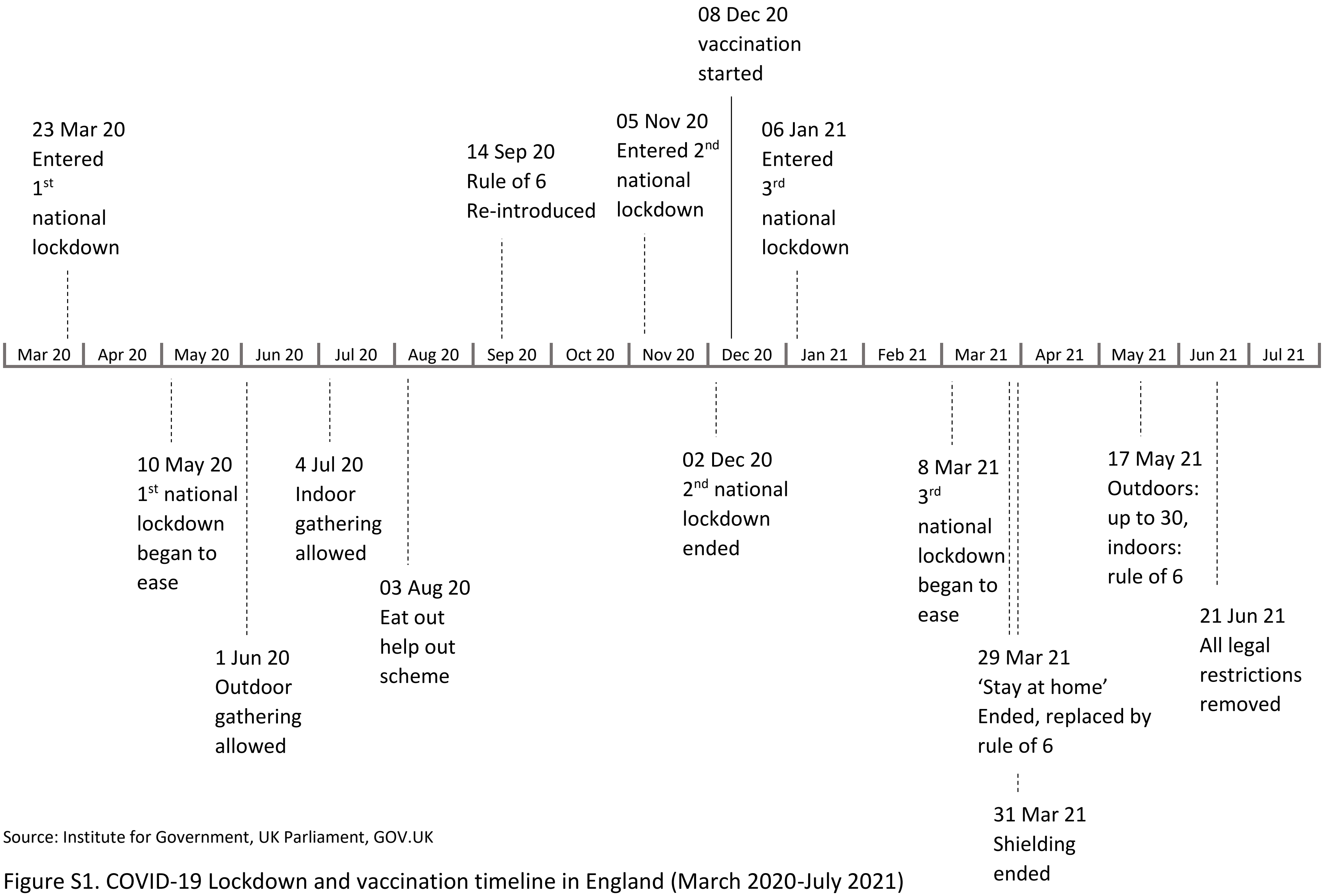


Source: Institute for Government, UK Parliament, GOV.UK

Figure S1. COVID-19 Lockdown and vaccination timeline in England (March 2020-July 2021)

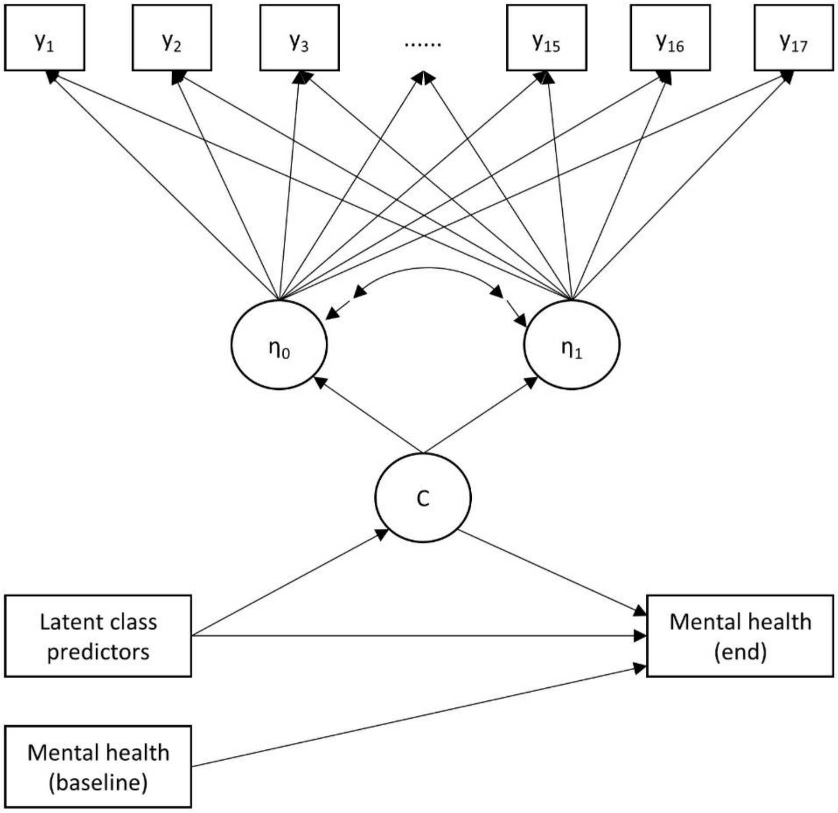


Figure S2. Full model: growth mixture model with covariates and distal outcomes

Table S1. Dates corresponding to different time points (months)

| Date | Month |
|---|---|
| 21/03/20-17/04/20 | 1 |
| 18/04/20-15/05/20 | 2 |
| 16/05/20-12/06/20 | 3 |
| 13/06/20-10/07/20 | 4 |
| 11/07/20-07/08/20 | 5 |
| 24/08/20-20/09/20 | 6 |
| 21/09/20-18/10/20 | 7 |
| 19/10/20-15/11/20 | 8 |
| 16/11/20-13/12/20 | 9 |
| 14/12/20-10/01/21 | 10 |
| 11/01/21-07/02/21 | 11 |
| 08/02/21-22/02/21 | 12 |
| 08/03/21-04/04/21 | 13 |
| 05/04/21-02/05/21 | 14 |
| 03/05/21-30/05/21 | 15 |
| 31/05/21-27/06/21 | 16 |
| 28/06/21-25/07/21 | 17 |

Table S2. Model fit indices for different model specifications

| Model specification | BIC | ABIC | LMR-LR | ALMR-LR | Entropy |
|---|---|---|---|---|---|
| 1-class GMM | 1,060,616 | 1,060,498 | NA | NA | NA |
| 2-class GMM | 1,057,617 | 1,057,490 | <0.001 | <0.001 | 0.689 |
| 3-class GMM | 1,055,055 | 1,054,918 | 0.003 | 0.004 | 0.657 |
| 4-class GMM | 1,054,380 | 1,054,234 | 0.290 | 0.299 | 0.718 |

Table S3. Results from the one-step condition GMM (N=25,390)

| | Adaptive (vs Home-confined) C2 (vs C1) | | Unconfined (vs Home-confined) C3 (vs. C1) | |
|---|---|---|---|---|
| | OR | 95% CI | OR | 95% CI |
| Women (vs. men) | **0.80** | **[0.66-0.97]** | **0.59** | **[0.51-0.70]** |
| Ethnic minority (vs. white) | 1.07 | [0.74-1.55] | **0.62** | **[0.44-0.89]** |
| Age: 30-45 (vs. 18-29) | 1.00 | [0.70-1.43] | 1.37 | [0.98-1.91] |
| Age: 46-59 (vs. 18-29) | **0.58** | **[0.41-0.84]** | 1.41 | [1.02-1.96] |
| Age: 60+ (vs. 18-29) | **0.59** | **[0.41-0.85]** | **1.74** | **[1.25-2.43]** |
| Education: A levels (vs. GCSEs or below) | 1.06 | [0.86-1.32] | 1.18 | [0.98-1.43] |
| Education: degree+ (vs. GCSEs or below) | 1.01 | [0.81-1.26] | **1.74** | **[1.45-2.09]** |
| Low income: <30k (vs. ≥30k) | **0.79** | **[0.65-0.97]** | **0.61** | **[0.52-0.72]** |
| Employed (vs. other) | **2.62** | **[2.13-3.22]** | **2.64** | **[2.19-3.17]** |
| Rural (vs. urban) | 1.23 | [1.00-1.51] | 0.96 | [0.81-1.14] |
| Own a dog (vs. none) | 0.99 | [0.79-1.25] | **2.84** | **[2.37-3.41]** |
| Living alone (vs with others) | **0.66** | **[0.52-0.82]** | 0.94 | [0.78-1.13] |
| Number of close friends | **1.05** | **[1.01-1.08]** | **1.06** | **[1.03-1.10]** |
| Frequency of social contacts | **1.40** | **[1.29-1.51]** | **1.45** | **[1.35-1.56]** |
| Physical health diagnosis (vs. no diagnosis) | **0.65** | **[0.53-0.79]** | **0.34** | **[0.29-0.40]** |
| Mental health diagnosis (vs. no diagnosis) | **0.60** | **[0.48-0.76]** | **0.57** | **[0.47-0.69]** |
| Personality: neuroticism | 1.03 | [1.00-1.05] | 1.02 | [1.00-1.04] |
| Personality: extraversion | **1.05** | **[1.02-1.07]** | **1.03** | **[1.01-1.05]** |
| Personality: openness | 0.98 | [0.95-1.00] | **0.96** | **[0.94-0.99]** |
| Personality: agreeableness | 0.99 | [0.96-1.02] | **0.96** | **[0.94-0.99]** |
| Personality: conscientiousness | 1.03 | [1.00-1.07] | **1.08** | **[1.06-1.11]** |
| COVID-19 stress minor (vs. none) | 1.19 | [0.96-1.46] | 0.92 | [0.78-1.09] |
| COVID-19 stress major (vs. none) | 0.95 | [0.76-1.19] | **0.57** | **[0.47-0.69]** |
